# Evaluating the Efficacy of COVID-19 Vaccines

**DOI:** 10.1101/2020.10.02.20205906

**Authors:** Dan-Yu Lin, Donglin Zeng, Devan V. Mehrotra, Lawrence Corey, Peter B. Gilbert

## Abstract

A large number of studies are being conducted to evaluate the efficacy and safety of candidate vaccines against novel coronavirus disease-2019 (COVID-19). Most Phase 3 trials have adopted virologically confirmed symptomatic COVID-19 disease as the primary efficacy endpoint, although laboratory-confirmed SARS-CoV-2 is also of interest. In addition, it is important to evaluate the effect of vaccination on disease severity. To provide a full picture of vaccine efficacy and make efficient use of available data, we propose using SARS-CoV-2 infection, symptomatic COVID-19, and severe COVID-19 as dual or triple primary endpoints. We demonstrate the advantages of this strategy through realistic simulation studies. Finally, we show how this approach can provide rigorous interim monitoring of the trials and efficient assessment of the durability of vaccine efficacy.

**Summary:** To increase statistical power and meet vaccine success criteria, we propose to evaluate the efficacy of COVID-19 vaccines by using the dual or triple primary endpoints of SARS-CoV-2 infection, symptomatic COVID-19, and severe COVID-19.

## Introduction

There is an urgent need to develop effective vaccines against SARS-CoV-2, the virus causing the global COVID-19 pandemic. Several candidate vaccines have shown strong immune responses and acceptable safety profiles and have moved rapidly into large-scale Phase 3 trials.^1–8^ As of December 8, 2020, a total of 28 Phase 3 trials on 13 candidate vaccines have been launched around the world.^7^ Through Operation Warp Speed, the US government has selected several of these candidates for Phase 3 testing, including mRNA vaccines encoding the prefusion stabilized SARS-CoV-2 Spike protein (mRNA-1273, BNT162b1)^2,3^, a recombinant replication-defective chimpanzee adenovirus expressing a wild-type SARS-CoV-2 Spike protein (AZD1222)^4^, a recombinant, replication-incompetent adenovirus type 26 (Ad26) vector vaccine encoding a stabilized SARS-CoV-2 Spike protein (Ad26.COV2.S)^5^, a SARS-CoV-2 recombinant stabilized Spike protein vaccine with AS03 adjuvant, and a SARS-CoV-2 recombinant stabilized Spike protein nanoparticle vaccine (SARS-CoV-2 rS) with Matrix-M1™ adjuvant.^6^

The vaccine regimens have generally protected against COVID-19 disease endpoints in animal models^5^ and have induced binding and neutralizing antibody responses to vaccine-insert Spike proteins in most vaccine recipients, exceeding response levels seen in convalescent sera.^2–4,6^ The antibody marker endpoints are of the types that have been accepted as surrogate endpoints for many approved vaccines^9^, generating enthusiasm that the vaccines can plausibly confer protection. Interim results from Pfizer/BioNTech, Moderna, and AstraZeneca/Oxford University suggested high vaccine efficacy against COVID-19 disease.

Rapid introduction of effective vaccines in the US and other countries with high numbers of COVID-19 cases would be a major step toward halting the global pandemic. However, deployment of a non-effective vaccine could actually worsen the pandemic because public acceptance of a COVID-19 vaccine might diminish the implementation of other control measures. Thus, we need speedy and reliable evaluation of the efficacy of COVID-19 vaccines on the basis of clinically relevant endpoints.

Most Phase 3 trials have adopted virologically confirmed symptomatic COVID-19 illness as the primary efficacy endpoint, although laboratory-confirmed SARS-CoV-2 is also acceptable.^10^ It is possible that a vaccine is much more effective in preventing severe than mild COVID-19. Thus, we should also evaluate the effect of vaccination on severe COVID-19.^10^ However, a large sample size is likely required for a trial using a severe COVID-19 endpoint.

We propose using SARS-CoV-2 infection, symptomatic COVID-19, and severe COVID-19 as triple primary endpoints or using SARS-CoV-2 infection and symptomatic COVID-19 or symptomatic COVID-19 and severe COVID-19 as dual primary endpoints, the specific choice depending on the expected incidence of the three events and on the targeted vaccine efficacy for the three endpoints. This approach incorporates more evidence on vaccine efficacy into decision making than using only one of the three events as the primary endpoint. It can improve statistical power and increase the likelihood of meeting vaccine success criteria, thus accelerating the discovery and licensure of effective vaccines.

## Methods

We consider the endpoints of SARS-CoV-2 infection, symptomatic COVID-19, and severe COVID-19, referring to them as infection, disease, and severe disease, respectively. Suppose that a large number of individuals are randomly assigned to vaccine or placebo and that the trial records whether or not each participant has developed each of the three endpoints by the end of follow-up, as well as their length of follow-up.

We formulate the effect of the vaccine on each of the three endpoints through a Poisson model. Although investigators are mainly interested in the first occurrence of each event, the Poisson modeling approach provides a reasonable approximation to the data because the event rates for all three endpoints are relatively low. We define the vaccine efficacy in terms of the proportionate reduction in the event rate between vaccinated and un-vaccinated individuals.

The criteria for claiming that a vaccine is successful should be strict enough to ensure worthwhile efficacy. A vaccine whose efficacy is higher than 50% can markedly reduce incidence of COVID-19 among vaccinated individuals and help to build herd immunity. An advisory panel convened by the World Health Organization (WHO) recommended 50% vaccine efficacy for at least 6 months post vaccination as a minimal criterion to define an efficacious vaccine.^11^ The US Food and Drug Administration (FDA) guidance defines vaccine success criteria as a point estimate of vaccine efficacy at least 50% and the interim-monitoring adjusted lower bound of the 95% confidence interval exceeding 30%.^10^ The FDA guidance criteria do not specify a minimum period of follow-up. However, given the intent of current vaccine development to identify efficacious vaccines within several months of trial initiation, the expectation seems to be reliable evidence for vaccine efficacy over approximately 6 months, consistent with the WHO recommendation.

Many Phase 3 trials specify assessment of vaccine efficacy over longer-term follow-up as an important study objective. The FDA guidance document states that “A lower bound ≤ 30% but > 0% may be acceptable as a statistical success criterion for a secondary efficacy endpoint, provided that secondary endpoint hypothesis testing is dependent on the success on the primary endpoint.” This statement refers to earlier FDA guidance on a fixed-sequence testing method,^12^ under which vaccine efficacy is tested against a sequence of secondary endpoints in a pre-defined order, where tests of each endpoint are performed at the same significance level (one-sided type I error of 2.5%), moving to the next endpoint only after a success on the previous endpoint. The WHO Solidarity Trial protocol^13^ specifies symptomatic COVID-19 through longer term follow-up (ideally 12 months or more) and severe COVID-19 over the same time frame as secondary endpoints. Following these guidelines and precedents, we consider hypothesis testing of vaccine efficacy over 12 months as a secondary analysis, using a null hypothesis that is less stringent than the 30% null hypothesis value used for the primary analysis, recognizing that it is more difficult for a vaccine to provide 12-month than 6-month protection and that even moderate vaccine efficacy through 12 months could be an important characteristic of a COVID-19 vaccine. In sum, we consider both the assessment of vaccine efficacy against primary endpoints over six months, using a 30% null hypothesis, and the assessment of vaccine efficacy against the same endpoints over 12 months, using a 0% or 15% null hypothesis.

For each of the three endpoints, we obtain the maximum likelihood estimator for the vaccine efficacy under the Poisson model. In addition, we calculate the score statistic for testing the null hypothesis that the vaccine efficacy is less than a certain lower limit, say 30%, against the alternative hypothesis that the vaccine efficacy is greater than the lower limit; we divide the score statistic by its standard error to create a standard-normal test statistic.

We propose to test all three null hypotheses, adjusting the significance threshold for the three test statistics to control the overall type I error at the desired level. We consider a vaccine to be successful if any of the three null hypotheses is rejected. We describe this multiple testing method in greater detail in Supplemental Appendix 1, where we also describe a sequential testing procedure to determine which of the three null hypotheses should be rejected.

In the sequential testing procedure, we order the three hypotheses according to the order of the three observed test statistics, from the most extreme observed value to the least extreme. We test the first null hypothesis using the significance threshold from the aforementioned multiple testing procedure. If the first null hypothesis is rejected, we test the second null hypothesis by applying the multiple testing procedure to the remaining two test statistics. If the second null hypothesis is rejected, we test the last null hypothesis by using the unadjusted significance threshold.

Clearly, this sequential testing procedure is more powerful than the multiple testing procedure in identifying which endpoints the vaccine is efficacious against. Both the proposed multiple testing and sequential testing methods properly account for the correlations of the test statistics and thus are more powerful than the conventional Bonferroni correction and related multiplicity adjustments that assume independence of tests.

If the effects of a vaccine are expected to be similar among the three endpoints, then we can enhance statistical power by combining the evidence of the vaccine effects on the three endpoints and performing a single test of overall vaccine efficacy. Specifically, we propose taking the sum of the three score statistics and dividing the sum by its standard error to create a standard-normal test statistic. We refer to this method as the combined test (Supplemental Appendix 1); this is in the same vein as combining estimators for a common effect in meta-analysis.^14^

Instead of the triple primary endpoints, we may consider the dual primary endpoints of infection and disease if severe disease is very rare or the dual primary endpoints of disease and severe disease if the vaccine is expected to be only weakly effective against infection. Clearly, the above methods can be modified to test only two of the three endpoints.

It is desirable to periodically examine the accumulating data from a Phase 3 trial, so that the trial can be terminated if sufficient evidence emerges for a highly effective vaccine or a weakly effective candidate. In order to obtain rigorous stopping boundaries for a trial, we need to derive the joint distribution of the test statistics over interim looks. In Supplemental Appendix 2, we show that the proposed test statistics over interim looks are jointly normal with the independent increment structure, such that standard methods for interim analyses^15–18^ can be applied.

## Results

We first conducted a series of simulation studies to compare the performance of the proposed methods with the use of a single primary endpoint in evaluating short-term vaccine efficacy. We assigned 27,000 subjects to vaccine or placebo at a ratio of 1:1. We assumed that subjects were enrolled at a constant rate over a 2-month period and vaccine efficacy was evaluated 6 months after the first subject was enrolled. We let 1% of the placebo subjects to acquire infection, 0.6% to experience disease, and 0.12% to develop severe disease (Supplemental Appendix 3). These event proportions were based on the assumption of annualized incidence of about 1.5% for symptomatic COVID-19 disease in the placebo group, together with the assumptions that about 40% of infections are asymptomatic and that about 20% of symptomatic COVID-19 cases will be severe. We set the vaccine efficacy for disease, denoted by VE_*D*_, to 60%; we set the vaccine efficacy for infection, denoted by VE_*I*_, to 40%, 50%, 55% or 60%; and we set the vaccine efficacy for severe disease, denoted by VE_*S*_, to 60%, 70%, 80% or 90% (Supplemental Appendix 3). For each combination of VE_*I*_, VE_*D*_, and VE_*S*_, we simulated 100,000 datasets. (The average number of each endpoint can be easily calculated. For example, there are approximately 189 cases of infection, 113 cases of disease, and 23 cases of severe disease under VE_*I*_ = VE_*D*_ = VE_*S*_ = 0.6.) In each data set, we tested the null hypothesis that the vaccine efficacy is at most 30% against the alternative hypothesis that the vaccine efficacy is greater than 30% at the one-sided nominal significance level of 2.5%. Table 1 summarizes the power of various methods for testing the null hypothesis of no worthwhile efficacy (i.e., at most 30%). Using the single endpoint of disease has 80% power under VE_*D*_ = 60%. Indeed, we chose the sample size and disease rate in the placebo group to achieve this power, which is considered the benchmark for other methods. When VE_*I*_ is equal to or slightly below VE_*D*_, the single endpoint of infection is more powerful than the single endpoint of disease (e.g., 96% versus 80% power under VE_*I*_ = VE_*D*_ = 60%) because infection is more frequent than disease. Due to low incidence, the single endpoint of severe disease has poor power unless VE_*S*_ is very high (e.g., 69% and 91% power under VE_*S*_ = 80% and 90%, respectively). The combined test for the dual endpoints of infection and disease and the combined test for the triple endpoints are substantially more powerful than using disease as the single endpoint when VE_*I*_ is similar to VE_*D*_ (e.g., 94% and 93% power for the two combined tests versus 80% power for the single endpoint of disease under VE_*I*_ = VE_*D*_ = VE_*S*_ = 60%). The combined test for the dual endpoints of disease and severe disease is more powerful than the single endpoint of disease when VE_*S*_ is high (e.g., 93% versus 80% power under VE_*S*_ = 90%). The combined test is more powerful than multiple testing for the dual endpoints of disease and severe disease, but the opposite is true for the dual endpoints of infection and disease and the triple primary endpoints when VE_*I*_ is low. The proposed multiple-testing method is appreciably more powerful than Bonferroni correction.

**Table 1.** Statistical Power (%) for Testing the Null Hypothesis of At Most 30% Vaccine Efficacy Against Infection (I), Disease (D), and Severe Disease (S) Over 6 Months

| Vaccine Efficacy |  |  | Single Endpoint |  |  | Combined Test |  |  | Multiple Testing |  |  | Bonferroni |  |  |
| --- | --- | --- | --- | --- | --- | --- | --- | --- | --- | --- | --- | --- | --- | --- |
| $VE_I$ | $VE_D$ | $VE_S$ | I | D | S | I-D | D-S | I-D-S | I-D | D-S | I-D-S | I-D | D-S | I-D-S |
| 40% | 60% | 60% | 21 | 80 | 27 | 51 | 77 | 53 | 75 | 75 | 72 | 73 | 74 | 69 |
| 40% | 60% | 70% | 21 | 80 | 45 | 51 | 83 | 57 | 75 | 78 | 75 | 73 | 77 | 72 |
| 40% | 60% | 80% | 21 | 80 | 69 | 51 | 88 | 61 | 75 | 85 | 82 | 73 | 84 | 79 |
| 40% | 60% | 90% | 21 | 80 | 91 | 51 | 93 | 65 | 75 | 94 | 92 | 73 | 93 | 90 |
| 50% | 60% | 60% | 65 | 80 | 27 | 78 | 77 | 78 | 79 | 75 | 76 | 77 | 74 | 73 |
| 50% | 60% | 70% | 65 | 80 | 45 | 78 | 83 | 81 | 79 | 78 | 78 | 77 | 77 | 75 |
| 50% | 60% | 80% | 65 | 80 | 69 | 78 | 88 | 83 | 79 | 85 | 84 | 77 | 84 | 81 |
| 50% | 60% | 90% | 65 | 80 | 91 | 78 | 93 | 86 | 79 | 94 | 93 | 77 | 93 | 91 |
| 55% | 60% | 60% | 84 | 80 | 27 | 87 | 77 | 86 | 86 | 75 | 83 | 84 | 74 | 80 |
| 55% | 60% | 70% | 84 | 80 | 45 | 87 | 83 | 89 | 86 | 78 | 84 | 84 | 77 | 82 |
| 55% | 60% | 80% | 84 | 80 | 69 | 87 | 88 | 91 | 86 | 85 | 88 | 84 | 84 | 87 |
| 55% | 60% | 90% | 84 | 80 | 91 | 87 | 93 | 93 | 86 | 94 | 95 | 84 | 93 | 94 |
| 60% | 60% | 60% | 96 | 80 | 27 | 94 | 77 | 93 | 94 | 75 | 92 | 93 | 74 | 91 |
| 60% | 60% | 70% | 96 | 80 | 45 | 94 | 83 | 94 | 94 | 78 | 93 | 93 | 77 | 92 |
| 60% | 60% | 80% | 96 | 80 | 69 | 94 | 88 | 96 | 94 | 85 | 95 | 93 | 84 | 94 |
| 60% | 60% | 90% | 96 | 80 | 91 | 94 | 93 | 97 | 94 | 94 | 98 | 93 | 93 | 97 |

Note: $VE_I$ , $VE_D$ , and $VE_S$ denote, respectively, the vaccine efficacy for infection, disease, and severe disease. I-D, D-S, and I-D-S denote, respectively, the dual endpoints of infection and disease, the dual endpoints of disease and severe disease, and the triple endpoints of infection, disease, and severe disease. The power pertains to a single test at the one-sided nominal significance level of 2.5%.

In order to investigate the ability of the proposed methods in detecting long-term vaccine efficacy, we extended the follow-up time in the above simulation studies from a maximum of 6 months to a maximum of 12 months. We assumed that the event proportions for infection, disease, and severe disease in the placebo group over the 12-month period doubled those of the 6-month period. We reduced all values of vaccine efficacy by 30% to reflect the waning of vaccine efficacy against each endpoint over time. We tested the null hypothesis that the vaccine efficacy is 0% versus the alternative hypothesis that the vaccine efficacy is greater than 0% at the nominal significance level of 2.5%. The results are summarized in Table 2. Again, the proposed methods can substantially improve statistical power.

**Table 2.** Statistical Power (%) for Testing the Null Hypothesis of No Vaccine Efficacy Against Infection (I), Disease (D), and Severe Disease (S) Over 12 Months

| Vaccine Efficacy |  |  | Single Endpoint |  |  | Combined Test |  |  | Multiple Testing |  |  | Bonferroni |  |  |
| --- | --- | --- | --- | --- | --- | --- | --- | --- | --- | --- | --- | --- | --- | --- |
| $VE_I$ | $VE_D$ | $VE_S$ | I | D | S | I-D | D-S | I-D-S | I-D | D-S | I-D-S | I-D | D-S | I-D-S |
| 10% | 30% | 30% | 22 | 84 | 26 | 54 | 80 | 57 | 79 | 78 | 75 | 76 | 77 | 72 |
| 10% | 30% | 40% | 22 | 84 | 44 | 54 | 85 | 60 | 79 | 80 | 77 | 76 | 79 | 74 |
| 10% | 30% | 50% | 22 | 84 | 65 | 54 | 89 | 64 | 79 | 84 | 81 | 76 | 83 | 79 |
| 10% | 30% | 60% | 22 | 84 | 84 | 54 | 92 | 67 | 79 | 90 | 88 | 76 | 89 | 86 |
| 20% | 30% | 30% | 70 | 84 | 26 | 82 | 80 | 81 | 83 | 78 | 78 | 80 | 77 | 76 |
| 20% | 30% | 40% | 70 | 84 | 44 | 82 | 85 | 84 | 83 | 80 | 80 | 80 | 79 | 78 |
| 20% | 30% | 50% | 70 | 84 | 65 | 82 | 89 | 86 | 83 | 84 | 84 | 80 | 83 | 82 |
| 20% | 30% | 60% | 70 | 84 | 84 | 82 | 92 | 88 | 83 | 90 | 90 | 80 | 89 | 88 |
| 25% | 30% | 30% | 88 | 84 | 26 | 90 | 80 | 89 | 89 | 78 | 86 | 87 | 77 | 84 |
| 25% | 30% | 40% | 88 | 84 | 44 | 90 | 85 | 91 | 89 | 80 | 87 | 87 | 79 | 85 |
| 25% | 30% | 50% | 88 | 84 | 65 | 90 | 89 | 93 | 89 | 84 | 89 | 87 | 83 | 88 |
| 25% | 30% | 60% | 88 | 84 | 84 | 90 | 92 | 94 | 89 | 90 | 93 | 87 | 89 | 92 |
| 30% | 30% | 30% | 97 | 84 | 26 | 96 | 80 | 95 | 96 | 78 | 94 | 95 | 77 | 93 |
| 30% | 30% | 40% | 97 | 84 | 44 | 96 | 85 | 96 | 96 | 80 | 95 | 95 | 79 | 94 |
| 30% | 30% | 50% | 97 | 84 | 65 | 96 | 89 | 97 | 96 | 84 | 96 | 95 | 83 | 95 |
| 30% | 30% | 60% | 97 | 84 | 84 | 96 | 92 | 97 | 96 | 90 | 97 | 95 | 89 | 96 |

Note: $VE_I$ , $VE_D$ , and $VE_S$ denote, respectively, the vaccine efficacy for infection, disease, and severe disease. I-D, D-S, and I-D-S denote, respectively, the dual endpoints of infection and disease, the dual endpoints of disease and severe disease, and the triple endpoints of infection, disease, and severe disease. The power pertains to a single test at the one-sided nominal significance level of 2.5%.

## Discussion

We have presented a simple and rigorous framework to consider the totality of evidence when evaluating the benefit of a COVID-19 vaccine in reducing SARS-CoV-2 infection, symptomatic COVID-19, and severe COVID-19. The proposed methods are more robust to different scenarios of vaccine efficacy than the use of a single primary endpoint. We recommend using the combined test to provide an overall assessment of worthwhile vaccine efficacy, then using the sequential test (Supplemental Appendix 1) to determine the endpoints against which the vaccine is efficacious.

If a vaccine is more effective in preventing severe than mild COVID-19, then using symptomatic COVID-19 and severe COVID-19 as dual primary endpoints will be more powerful than using either of the two events as a single primary endpoint. If the vaccine efficacy for infection is nearly as high as that for disease, then using infection, symptomatic COVID-19, and severe COVID-19 as triple primary endpoints will be the most powerful.

Most Phase 3 trials have targeted 90% power for detecting 60% (short-term) vaccine efficacy against COVID-19 disease. The actual power may be lower if the vaccine is less effective, the disease incidence is lower than anticipated, or it is an interim analysis. In our simulation studies, using disease as a single primary endpoint had only 80% power. However, the proposed methods could boost the power to 90%.

The Phase 3 trials under Operation Warp Speed have thus far used symptomatic COVID-19 as the sole primary endpoint, assessing severe COVID-19 as a secondary endpoint and assessing a composite burden-of-disease endpoint as either a secondary endpoint or an exploratory endpoint.^19^ Under such a plan with a fixed-sequence strategy, hypothesis testing on secondary endpoints would be permitted only if the result on the primary endpoint is statistically significant.^12^ In the likely scenarios that VE_*S*_ is higher than VE_*D*_, using disease and severe disease as dual primary endpoints will be more powerful than using disease alone as the sole primary endpoint and thus may accelerate the discovery and deployment of effective vaccines.

We have focused on vaccine trials for populations enriched with high-risk individuals (e.g., front-line health-care personnel, factory workers, older adults, people with underlying health conditions), in which the risks for infection, disease, and severe disease are all appreciable. In generally healthy populations, such as college students, the majority of infections are asymptomatic, and severe disease is rare. For such settings, power can be maximized by using the dual primary endpoints of infection and disease.

We have used Poisson models instead of Cox proportional hazards models for several reasons. First, there are considerable inaccuracies in determining the event times, especially the infection time; the Poisson modeling approach requires only the knowledge of whether or not the event has occurred by the end of follow-up. Second, Poisson models are simpler than Cox models, both conceptually and computationally. Because the event rates are relatively low, the two modeling approaches should provide similar results.^20^ We fitted both Poisson and Cox models in our simulation studies, and the power of the two approaches was nearly identical (Supplemental Appendix 3).

We have emphasized hypothesis testing based on score statistics. In Supplemental Appendix 4, we extend our work to general Poisson regression, which can be used to estimate vaccine efficacy, construct confidence intervals, compare multiple vaccines, and accommodate baseline risk factors (e.g., age, gender, race, occupation, co-morbidity). Baseline risk factors can have major impact on the occurrences of SARS-CoV-2 infection, symptomatic COVID-19, and severe COVID-19. In addition, some participants in COVID-19 vaccine efficacy trials may become unblinded through the use of available diagnostic tests, and at some point trials may become unblinded. Covariate adjustment in the analysis of vaccine efficacy against endpoints during post unblinding follow-up is important for minimizing bias due to potential differences in exposure to SARS-CoV-2 between the vaccine and placebo arms.

We have developed our methods in order to accelerate the discovery, characterization, and licensure of effective COVID-19 vaccines. An important function of the Phase 3 trials is to continue the unblinded follow-up of the vaccine and placebo groups after definite evidence of short-term efficacy has emerged, so as to assess duration of protection and improve precision for assessment of prevention of severe disease, as well as for assessment of safety. Duration of vaccine efficacy is an influential parameter in models of population impact of deployed vaccines, and understanding of how vaccine efficacy wanes over time is essential to deciding whether or not booster vaccinations may be required and to estimating the optimal timing of the boosts. The ability of our framework to use the joint distribution of the estimators to provide more precise confidence intervals around the three vaccine efficacy parameters than existing methods (e.g., Bonferroni correction) that do not account for the correlation of endpoints is advantageous regardless of whether one, two, or three endpoints are selected as primary.

## Data Availability

The simulated datasets are available from the corresponding author upon reasonable request.

## Acknowledgments

The authors are grateful to Yu Gu and Bridget I. Lin for assistance and to two referees for constructive comments. This work was supported by the National Institutes of Health.

## Supplemental Appendix

### 1. Two-Group Comparisons

Suppose that we record the occurrences of *K* types of events (e.g., SARS-CoV-2 infection, symptomatic COVID-19, severe COVID-19) for a total of *n* subjects. For *k* = 1, …, *K* and *i* = 1, …, *n*, let *Y*_*ki*_ denote the number of the *k*th type of event experienced by the *i*th subject, and *T*_*ki*_ denote the corresponding follow-up time. For *i* = 1, …, *n*, let *X*_*i*_ indicate, by the values 1 versus 0, whether the *i*th subject receives vaccine or placebo. We assume that *Y*_*ki*_ follows a Poisson distribution with mean 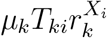, where *µ*_*k*_ is the event rate (per unit time interval) for placebo, and *r*_*k*_ is the rate ratio (i.e., relative risk) between vaccine and placebo. The vaccine efficacy on the *k*th type of event is defined by VE_*k*_ = 1 −*r*_*k*_, which is the proportionate reduction in cases among the vaccinated persons.

The likelihood for (*µ*_*k*_, *r*_*k*_) takes the form

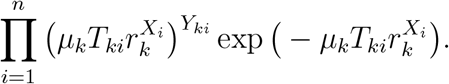

The maximum likelihood estimator of *r*_*k*_ is

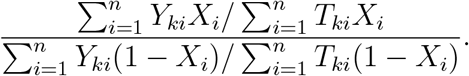

The score statistic for testing the null hypothesis *H*_*k*_ : *r*_*k*_ ≥ *r*_*k*0_, i.e., VE_*k*_ ≤ 1 − *r*_*k*0_, against the alternative hypothesis that *r*_*k*_ *< r*_*k*0_, i.e., VE_*k*_ > 1 − *r*_*k*0_, takes the form

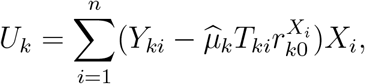

where 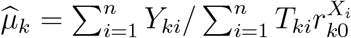. For large *n*, the vector of score statistics (*U*_1_, …, *U*_*K*_) is *K*-variate zero-mean normal with covariance matrix {*V*_*kl*_; *k, l* = 1, …, *K*}, where

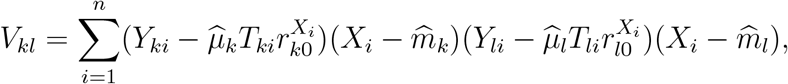

and 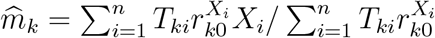.

For *k* = 1, …, *K*, we test the null hypothesis *H*_*k*_ : *r*_*k*_ ≥ *r*_*k*0_ by using the *Z*-score: 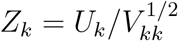, which is standard normal under the null hypothesis. We propose to test the *K* null hypotheses, adjusting the critical value so as to control the overall type I error at *α*. Specifically, we reject *H*_*k*_ if the observed value of *Z*_*k*_ is less than the constant *c* that satisfies the equation

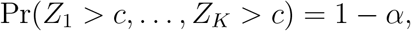

where (*Z*_1_, …, *Z*_*K*_) is zero-mean *K*-variate normal with covariance matrix {*ρ*_*kl*_; *k, l* = 1, …, *K*}, and *ρ*_*kl*_ = *V*_*kl*_*/*(*V*_*kk*_*V*_*ll*_)^1*/*2^. We refer to this method as multiple testing.

To determine which types of events the vaccine is effective against, we adopt a sequential testing procedure, which is more powerful than the above multiple testing method. Let 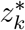 be the *k*th smallest observed value of the *Z*_*k*_’s, and let 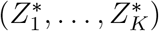 be a zero-mean *K*-variate normal random vector with a covariance matrix obtained by rearranging the rows and columns of {*ρ*_*kl*_; *k, l* = 1, …, *K*} according to the order 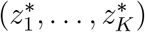. In addition, let 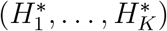 be the ordered sequence of the *H*_*k*_’s according to the order 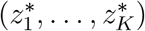. Starting with 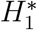, we reject 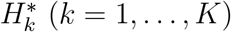 if

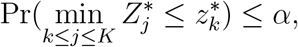

provided that 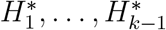 have been tested and rejected. It can be shown that the Type I error probability of this procedure is *α* for any combination of the true *H*_*k*_’s.^1^

We also propose to test the global null hypothesis of no worthwhile vaccine benefit on any endpoint, i.e., *H*_0_ : *r*_*k*_ ≥ *r*_*k*0_ for all *k* = 1, …, *K*, by combining the evidence of the vaccine effects on the *K* endpoints. Specifically, we form a new test statistic by summing the *K* score statistics and dividing the sum by its standard error:

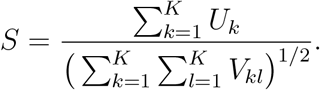

We refer to *S* as the combined score test, which is standard normal under *H*_0_.

### 2. Interim Analyses

Suppose that we perform interim analyses at times *t*_1_ *< t*_2_ *<* … *< t*_*M*_. Let *U*_*k*_(*t*) be the score statistic *U*_*k*_ based on the data collected up to time *t*. Write 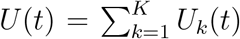. By the multivariate central limit theorem, the random vector {*U* (*t*_1_), …, *U* (*t*_*M*_)} is *M*-variate normal. Under the Poisson assumption, for *s > t*, the covariance between *U*_*k*_(*s*) and *U*_*l*_(*t*) is equal to the covariance between *U*_*k*_(*t*) and *U*_*l*_(*t*). If follows that the covariance between *U* (*s*) and *U* (*t*) is equal to the variance of *U* (*t*). Because of this independent increment property, all standard methods for interim analyses, such as group sequential tests and stochastic curtailment,^2–5^ are applicable to the combined test, as well as the individual tests.

### 3. Simulation Studies

We assigned 27,000 subjects to vaccine or placebo at a ratio of 1:1. For each subject in the placebo group, we generated the time from randomization to infection, the time from infection to disease, and the time from disease to severe disease from the exponential distributions (Fig. 1) with means *ξλ*_1_, *ξλ*_2_, and *ξλ*_3_, respectively, where *ξ* is a subject-specific random effect that has a gamma distribution with mean 1 and variance 0.5. In the first set of simulation studies, we generated the follow-up time from the Uniform (120,180) distribution. We chose *λ*_1_, *λ*_2_, and *λ*_3_ to yield event proportions of 1% for infection, 0.6% for disease, and 0.12% for severe disease over the 6-month follow-up period. For each subject in the vaccine group, we generated the three event times and the follow-up time in the above manner but chose *λ*_1_, *λ*_2_, and *λ*_3_ to yield the desired values of VE_*I*_, VE_*D*_, and VE_*S*_. To assess statistical power, we set VE_*D*_ to 0.6, VE_*I*_ to 0.4, 0.5, 0.55 or 0.6, and VE_*S*_ to 0.6, 0.7, 0.8 or 0.9. In the second set of simulation studies, we generated the follow-up time from the Uniform (300,360) distribution and chose the event proportions of 2% for infection, 1.2% for disease, and 0.24% for severe disease over the 12-month follow-up period. We set VE_*D*_ to 0.3, VE_*I*_ to 0.1, 0.2, 0.25 or 0.3, and VE_*S*_ to 0.3, 0.4, 0.5 or 0.6.

**Figure 1.**
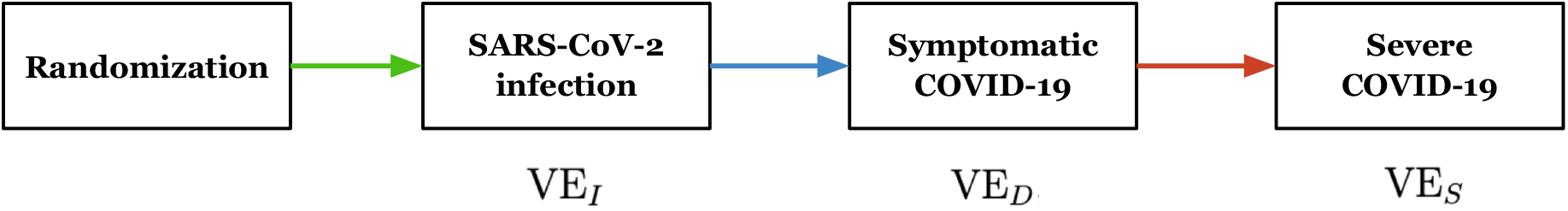
A 4-state model for a Phase 3 COVID-19 vaccine trial. The time between two adjacent events follows an exponential distribution, with different rates between vaccine and placebo to achieve vaccine efficacy of VE_*I*_, VE_*D*_, and VE_*S*_ for infection, disease, and severe disease, respectively.

We evaluated a total of 12 methods: (1) *Z*_1_ alone; (2) *Z*_2_ alone; (3) *Z*_3_ alone; (4) combining *U*_1_ and *U*_2_; (5) combining *U*_2_ and *U*_3_; (6) combining *U*_1_, *U*_2_, and *U*_3_; (7) multiple testing with *Z*_1_ and *Z*_2_; (8) multiple testing with *Z*_2_ and *Z*_3_; (9) multiple testing with *Z*_1_, *Z*_2_, and *Z*_3_; (10) Bonferroni correction for *Z*_1_ and *Z*_2_; (11) Bonferroni correction for *Z*_2_ and *Z*_3_; and (12) Bonferroni correction for *Z*_1_, *Z*_2_, and *Z*_3_. For each method, we performed a one-sided test with the nominal significance level of 2.5% for the null hypothesis that the vaccine efficacy is at most 0.3 in the first set of simulation studies and is at most zero in the second set of simulation studies. We estimated the power for the 12 methods by simulating 100,000 datasets for each of the 16 combinations of VE_*I*_, VE_*D*_, and VE_*S*_. We considered *Z*_2_ alone the benchmark since most vaccine trials have adopted symptomatic COVID-19 as the primary endpoint.

We implemented the Poisson regression approach described in the previous section, as well as a Cox regression analog.^1,6^ The results of the two approaches are almost identical. Here, we report only the Poisson regression results. Poisson regression has clear advantages over Cox regression because it is computationally simple and does not require knowing the event time, but rather whether or not the subject has developed the event of interest by the end of follow-up.

### 4. General Regression

We consider general Poisson regression, which can be used to perform point and interval estimation, to compare multiple vaccines, and to accommodate baseline risk factors (e.g., age, gender, race, occupation, co-morbidities). As in Appendix 1, there are *K* types of events and a total *n* study subjects. For *k* = 1, …, *K* and *i* = 1, …, *n*, let *Y*_*ki*_ denote the number of the *k*th type of event experienced by the *i*th subject, *T*_*ki*_ denote the corresponding follow-up time, and *X*_*ki*_ denote a set of covariates (i.e., vaccine indicators, baseline risk factors). (We allow risk factors to depend on the event type.) We assume that *Y*_*ki*_ follows a Poisson distribution with mean 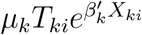, where *µ*_*k*_ is the baseline event rate (per unit time interval), and *β*_*k*_ is a set of log relative risks.

The likelihood for (*µ*_*k*_, *β*_*k*_) takes the form

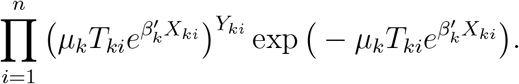

The profile-likelihood score function for *β*_*k*_ is

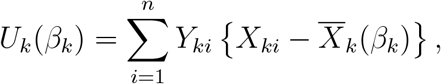

where 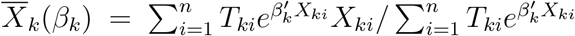. We obtain the maximum likelihood estimator for *β*_*k*_, denoted by 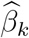, by solving the score equation *U*_*k*_(*β*_*k*_) = 0 via the Newton-Raphson algorithm. We then estimate 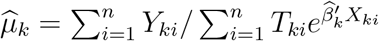.

For large *n*, the estimators 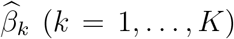 are jointly normal with means *β*_*k*_ (*k* = 1, …, *K*). In addition, the covariance matrix between 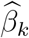 and 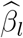 is 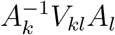, where 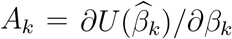, and

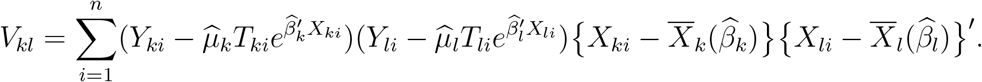

Let *η*_*k*_ be the component of *β*_*k*_ that corresponds to the vaccine effect on the *k*th endpoint. We extract the maximum likelihood estimator 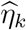 from 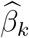 and extract the covariance matrix of 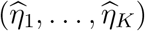, denoted by Σ = {*σ*_*kl*_; *k, l* = 1, …, *K*}, from the covariance matrices between 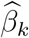and 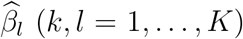. For *k* = 1, …, *K*, we calculate the *Z*-score for testing the null hypothesis *H*_*k*_ : *η*_*k*_ ≥ *η*_*k*0_ by using the *Z*-score: 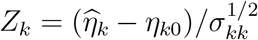. We can then use these *Z*-scores for the multiple testing and sequential testing procedures described in Appendix 1.

Suppose that *η*_1_ = *η*_2_ = … = *η*_*K*_ = *η*. Then we can estimate *η* by the weighted linear combination: 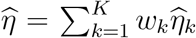, where (*w*_1_,…,*w*_*k*_)′ = (e′∑^-1^*e*)^-1^ ∑^-1^ *e*, and *e* = (1, …, 1)′.^1^ In addition, we test the global null hypothesis *H*_0_ : *η*_*k*_ ≥ *η*_*k*0_ for all *k* = 1, …, *K* by the standard-normal statistic:

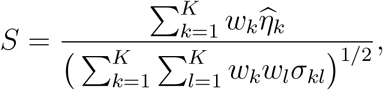

which is analogous to the combined score test given at the end of Appendix 1. Although the assumption of a common vaccine effect on the *K* endpoints may not hold, 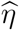 provides a concise summarization of the vaccine effects, and *S* provides a valid test of overall vaccine efficacy.

